# COVID-19 outbreak in Italy: estimation of reproduction numbers over two months toward the Phase 2

**DOI:** 10.1101/2020.05.12.20076794

**Authors:** Mattia Allieta, Andrea Allieta, Davide Rossi Sebastiano

**Author notes:** **Corresponding authors:** Davide Rossi Sebastiano, MD. PhD, **Email:**, Mattia Allieta, PhD, **Email:**. **Patients Involvement:** No patients were involved.

## Abstract

After two months from the first case in COVID-19 outbreak, Italy counts more than 190,000 confirmed positive cases. From the beginning of April 2020, the nationwide lockdown started to show early effects by reducing the total cumulative incidence reached by the epidemic wave. This allows the government to program the measures to loosen lockdown restrictions for the so called “Phase 2”. Here we provided the reproduction number estimation both in space and in time from February 24^th^ to April 24^th^, 2020 across two months into the epidemic. Our estimates suggest basic reproduction number averaged over all the regions of 3.29, confirming that epidemiological figures of the SARS-CoV-2 epidemic in Italy are higher than those observed at the early stage of Wuhan (China) outbreak. Based on the SARS-CoV-2 transmission dynamics reported here, we gave a quantitative evaluation of the efficiency of the government measures to low the reproduction number under the unity (control regime). We estimated that among the worst hit regions in Italy, Lombardy reached the control regime on March 22^nd^ followed by Emilia-Romagna (March 23^th^), Veneto (March 25^th^) and Piemonte (March 26^th^). Overall, we found that the mean value of time to reach the control regime in all the country is about 31 days from the February 24^th^ and about 14 days from the first day of nationwide lockdown (March 12^th^). Finally, we highlighted the interplay between the reproduction number and two demographic indices in order to probe the “state of activity” of the epidemic for each Italian region in the control regime. We believe that this approach can provide a tool in the management of “Phase 2”, potentially helping in challenging decision to continue, ease or tighten up restrictions.

## 1. Introduction

After the first COVID-19 case was diagnosed in Lombardy, Italy, on February 20th, 2020, [1] the novel coronavirus rapidly spread across the country leading to a dramatic spike in the number of new positive cases and deaths. To minimize the likelihood that people who were not infected come into contact with people who had contracted the disease, the Italian government imposed a series of progressively more strict social distancing measures which culminated in a national lock-down announced on March 11th, 2020. [2]

Around two months from the first case and more than 190,000 confirmed positive cases later, from the beginning of April, the effect of the nationwide lockdown started to achieve some level of success and the number of new infections began to smoothly decrease. These early signs of a slowdown of the COVID-19 pandemic in Italy provide a comforting picture of the outbreak’s stabilization which is driving the government to periodically review its lockdown measures in view of the so called “Phase 2”, i.e. the period during which citizens will have to live together with the virus as of all the industrial sector, including the non-essential economic activities, will start to reopen. However, since the regional differences in the number of new positive cases has been reported to be huge, with the Northern regions of Italy (namely Lombardia) being most affected, the establishment of the proper precautions to plan the “Phase 2” is a truly complicated task.

The planned restrictions and permissions that will be applied could thus vary from region to region. In this context, the systematical estimation of key epidemiological parameters, for each region can provide insight into the speed at which the disease had spread and will give a useful tool to figure out if a differential approach at the regional level on the measures to apply for “Phase 2” is feasible to keep down the transmission of SARS-COV2. At the beginning of epidemic and during the lockdown phase, Italian Government and the mainstream of the local and national mass media have been emphasized the relevance of the basic reproduction number (*R_0_*), i.e. the average number of secondary cases generated by a single primary case in a theoretically fully susceptible (100%) population, as the most important and informative parameter to monitor the epidemic trends. Obviously, *R*_0_ has an undoubted relevance since when *R*_0_ > 1 the infection may spread in the population and more *R*_0_ is large and deeper would be the interventions needed to control the epidemic. On the other hand, if *R*_0_ < 1, on average the infectious individual infects less than one person and the epidemic falls in a so called “control regime” where it will not be sustained, and it will die out. Nevertheless, *R*_0_ is not the only parameter that affect the impact and the spreading of the disease over a population which may largely result even from several demographic and epidemiological factors.

In this communication, we provided an estimation of the basic reproduction number *R*_0_ for all the Italian regions by the cumulative confirmed COVID-19 cases continuously updated and made public at the website of Dipartimento della Protezione Civile. [3] In addition, we estimated the time dependent reproduction number *R_t_*, which is the average number of secondary cases generated by an infectious individual at time *t*. We linked *R_t_* related to the last date of our period of observation (April 24th, 2020), with two demographic and epidemiologic indices in a simple three-dimensional array in order to highlight the “state of activity” of the epidemic for each Italian region. We provide a useful tool in the management of “Phase 2”, potentially helping in challenging decision to continue, ease or tighten up restrictions.

## 2. Material and Methods

### 2.1. Demographic and epidemiological data

The official demographic data of the resident population, the surface and the population density updated on January 1st, 2019, for each Italian region and Italy were taken from the Italian National Institute of Statistics (Istituto Nazionale di Statistica, ISTAT) and reported in Table S1 (Supporting Information). [4]

The official data of COVID-19 epidemic in Italy was taken from the task force of the Dipartimento della Protezione Civile. Cumulative data are available at various aggregation levels, namely national, regional and provincial and are accessible on Github. [3] Data for the analysis were considered from February 24th to April 24th, 2020.

In this period, we collected the daily cumulative number of confirmed positive cases (N), the number of “active” confirmed positive cases (NA), i.e., the number of infected people living not recovered from COVID-19, and the “density of infected people” (DA), calculated as NA/surface and expressed like the population density as number of persons/Km^2^, for each Italian region and Italy.

### 2.2 Estimation of the reproduction number *R*_0_ and estimation of time dependent reproduction number *R_t_*

To obtain the estimation of reproduction number we use the maximum likelihood estimation (ML) method which assumes that the number of secondary cases caused by an index case is Poisson distributed with an expected value R. Given then observation of (N_0_, N_1_,…, N_t_) incident cases over consecutive time units, R is estimated by maximizing the following log-likelihood function [5]:

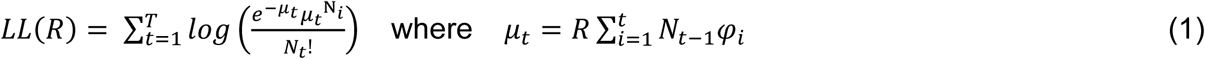

*φ_i_* is the distribution of the generation time corresponding to the distribution of the serial interval, i.e. the time between when a person gets infected and when they subsequently infect another other people, calculated at time *i* within the assumption that the incubation period does not change over the course of the epidemic [6]. We consider that the distribution of the serial interval was expected to follow a gamma distribution with mean (±SD) of 6.50 ± 4.03 days as reported by the Imperial College COVID-19 Response Team [7]. We note that this value agrees very well with gamma distribution with mean 6.6 days (95% CI, 0.7 to 19) recently determined from the analysis of 90 observations of individual serial intervals in 55 clusters in Lombardia (Italy) [8].

To estimate *R* = *R*_0_, the *LL(R)* function must be calculated over a period where epidemic curves showed exponential growth. As a first guess, to select this time window we used the simple procedure described by Obadia et al. [5] In brief, we computed the function over a range of possibile time periods by determining the deviance *r^2^* statistic for each iteration. Largest *r^2^* corresponds to the time window over which the ML model best described data.

To evaluate the time dependent reproduction number *R_t_* we adopted the method developed by Wallinga and Teunis. [9] The transmission probability (*p_ij_*) of individual *i* being infected by individual *j* at *t_i_, t_j_* onsets, respectively, can be described mathematically as:[5]

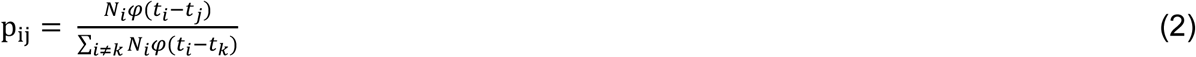

The net reproduction number *R_j_* is then then sum of all *p_ij_* involving *j* as the infector *R_j_ = ∑_j_p_ij_* and it can be averaged over all cases with same date of onset as 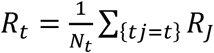.

Finally, since *R_t_* are computed by averaging over all transmission networks compatible with observed incidence data, no assumption is made about the time dependence of the epidemic unlike, for example the exponential growth in the well-known Bayesian approach. ^[5],[9]^ We believe, hence, that this model is particularly suitable to estimate the reproduction number in the post-peak period where the transmission is expected to decrease.

All the above data analyses were performed using the R0 package [5] as implemented in statistical software R. [10]

## 3. Results

### 3.1. Demographic and epidemiological data

Figure 1 shows COVID-19 incidence in Italy in the period of our observation, together with the dates in which Italian Government imposed restrictions, i.e. social distancing and school closure on March 4^th^, lockdown of Lombardia region and of 15 provinces in northern Italy on March 8^th^, national lockdown of Italy on March 11^th^. Overall, the incidence of COVID-19 infection in Italy shows that the exponential growth period may take place during the first 15-20 day from the national epidemic onset (February 24^th^, 2020).

**Figure 1.**
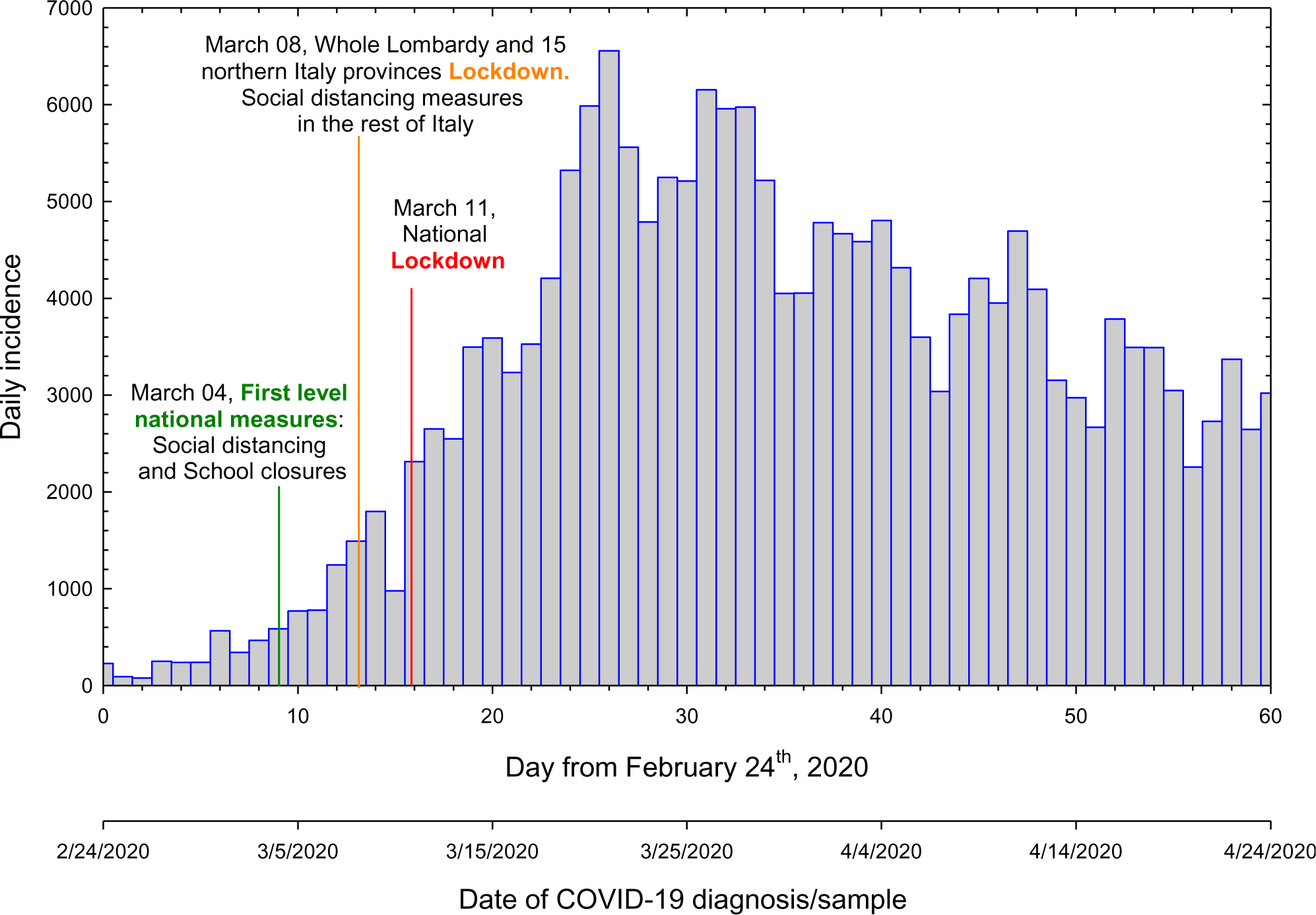
Daily number of new confirmed cases in Italy of the first two months of COVID-19 epidemic. Bars are incidence data of COVID-19 in Italy from February 24th to April 24th, 2020.[3]

Table S1-S2 (Supporting Information) show demographic and epidemiological data, respectively. As it is widely known, Table S2 shows that COVID-19 epidemic affected (and is affecting) harder the northern Italian regions, with N=16859 and NA=89384 on April 24th, i.e. more than 80% of the cases of the country (with 54,7% of the Italian resident population), if we aggregate epidemiological and demographic data of the northern regions (Lombardia, Piemonte, Veneto, Emilia Romagna, Liguria, Valle D’Aosta, Trentino-Alto Adige) plus Marche and Toscana regions. Furthermore, in Lombardia region epidemic had a huge spread, with N=71256 and NA= 34368 on April 24th, i.e. more than one third of the cases of the country (with 16.7% of the Italian resident population).

### 3.2. Estimation of the reproduction number R_0_ and estimation of time dependent reproduction number R_t_

In the top of the panels of Figures S1-S3 (Supporting Information) we reported the incidence data for all the regions plus Italy. Initial inspection of the datasets shows again that the exponential growth period may take place during the first 15-20 day from the relative epidemic onset. It should be noted that for the evaluation of *R*_0_ in the initial outbreak stage, we considered data from February 24^th^, up to March 18^th^, since data in a wider range can be affected by the national lockdown on March 11^th^.

In Figure 2(a) we showed *R*_0_ values obtained for SARS-COV-2 in all the regions and in Italy. Table 1 reports the same data represented in Figure 2(a), compared with those obtained by Riccardo *et al*., [2] D’Arienzo *et al*., [12] Distante *et al*. [13]. According to our ML estimation, the northern Italy regions Friuli Venezia Giulia *(R*_0_ = 3.61), Liguria *(R*_0_ = 3.68), Veneto *(R*_0_ = 3.73), Lombardia *(R*_0_ = 3.88), presented highest *R*_0_ suggesting that one infected person will infect up to four other people. The same tendency is shown in by the central region Lazio with *R*_0_ = 3.62. Apart the Southern regions Basilicata *(R*_0_ = 2.73), Molise *(R*_0_ = 2.52), Umbria *(R*_0_ = 2.44) showing the lowest values, the reproduction number *R*_0_ ranges from 3.00 to 3.49 in the rest of the country. The observed distribution of R0 related to all the country is reported in Figure 2(b).

**Figure 2.**
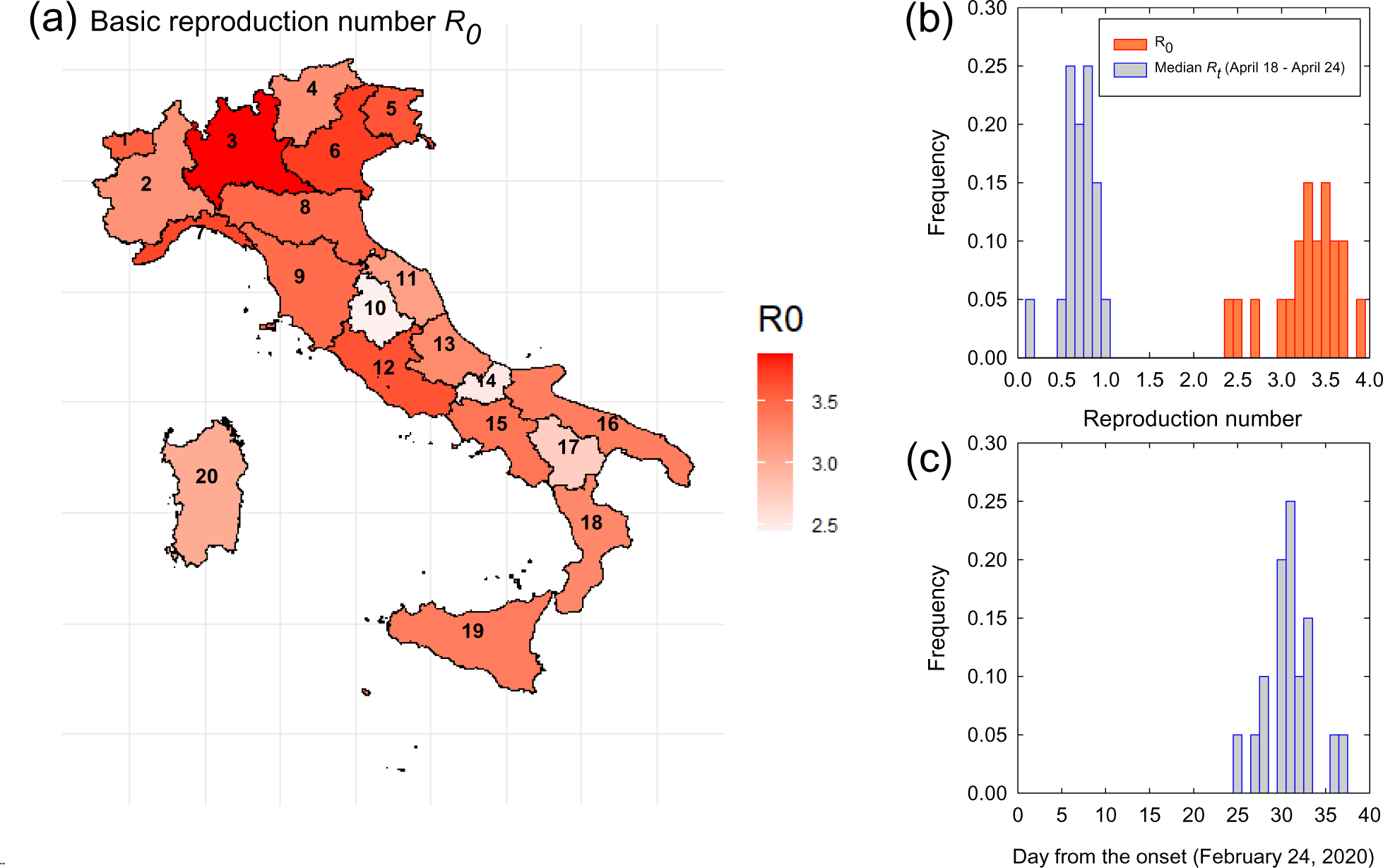
The basic reproduction number (*R_0_*) at the early stage of epidemic. (a) The map of Italy shows the basic reproduction number (R_0_) in all the regions as determined in the early stage of the epidemic. (b) Observed distribution of R_0_ and the R_t_ sorted by median values in the last 7-day time window (April 18^th^ – April 24^th^) determined in each region. (c) Observed distribution of the first day from the onset (February 24^th^) when the time evolution of R_t_ converges to ≲ 1 in each region. In panel (a) the different regions are numbered follows: 1 – Valle D'Aosta; 2 – Piemonte; 3 – Lombardia; 4 – Trentino-Alto Adige; 5 – Friuli-Venezia Giulia; 6 – Veneto; 7 – Liguria; 8 – Emilia-Romagna; 9 – Toscana; 10 – Umbria; 11 – Marche; 12 – Lazio; 13 – Abruzzo; 14 – Molise; 15 – Campania; 16 – Puglia; 17 – Basilicata; 18 – Calabria; 19 – Sicilia; 20 – Sardegna.

**Table 1.** Basic reproduction number *(R*_0_) values for SARS-CoV-2 estimated in our study and other three works. (*): The original incidence data related to Trento and Bolzano were merged into a single region called Trentino-Alto Adige resulting in a geographical disaggregation of Italy into 20 regions.

| Region | $R_0$ | | | |
| --- | --- | --- | --- | --- |
|  | This work | Riccardo et al.[2] | D'Arienzo et al. [11] | Distante et al.[12] |
| Abruzzo | 3.25 [95% CI: 2.82 to 3.73] | - | - | 3.03 |
| Basilicata | 2.73 [95% CI: 2.16 to 3.38] | - | - | 2.66 |
| Calabria | 3.27 [95% CI: 2.66 to 3.95] | - | - | 2.84 |
| Campania | 3.40 [95% CI: 3.01 to 3.82] | - | - | 3.14 |
| Emilia-Romagna | 3.49 [95% CI: 3.30 to 3.70] | 2.84 [95% CI: 2.57 to 3.13] | - | 3.38 |
| Friuli-Venezia Giulia | 3.61 [95% CI: 3.08 to 4.21] | - | - | 3.04 |
| Lazio | 3.62 [95% CI: 3.15 to 4.12] | 3.00 [95% CI: 2.68 to 3.33] | - | 3.11 |
| Liguria | 3.68 [95% CI: 3.29 to 4.09] | - | - | 3.10 |
| Lombardia | 3.88 [95% CI: 3.75 to 4.02] | 2.96 [95% CI: 2.73 to 3.17] | - | 3.60 |
| Marche | 3.09 [95% CI: 2.85 to 3.36] | - | - | 3.12 |
| Molise | 2.52 [95% CI: 1.68 to 3.60] | - | - | 2.60 |
| Piemonte | 3.19 [95% CI: 3.04 to 3.35] | - | - | 3.40 |
| Puglia | 3.34 [95% CI: 2.97 to 3.74] | 2.61 [95% CI: 2.13 – 3.13] | - | 3.11 |
| Sardegna | 3.00 [95% CI: 2.33 to 3.78] | - | - | 3.00 |
| Sicilia | 3.35 [95% CI: 2.95 to 3.78] | - | - | 2.99 |
| Toscana | 3.46 [95% CI: 3.12 to 3.83] | 2.50 [95% CI: 2.18 – 2.83] | - | 3.25 |
| Trentino-Alto Adige (*) | 3.21 [95% CI: 2.88 to 3.57] | - | - | Bolzano: 2.90<br>Trento: 3.23 |
| Umbria | 2.44 [95% CI: 2.16 to 2.74] | - | - | 2.79 |
| Valle D'Aosta | 3.43 [95% CI: 2.92 to 3.97] | - | - | 2.95 |
| Veneto | 3.73 [95% CI: 3.46 to 4.04] | 2.51 [95% CI: 2.18 to 2.86] | - | 3.32 |
| Italy | 3.22 [95% CI: 3.14 to 3.29] | - | 3.10 | - |

The time evolution of reproduction number *R_t_* for each region plus Italy is reported in the lower part of the panels of Figures S1-S3. All the *R_t_* profiles exhibited similar qualitative patterns characterized by large initial *R_t_* values followed by the decreasing over time of the parameter reaching *R_t_* < 1 close to the peak of daily incidence (~ 20-30 days). Below this value, *R_t_* further decreases featuring a sort of plateau in the *R_t_* < 1 “control regime” as detected for the country and for most of the regions. Uncertainty decreases over time because of the increased number of cases but at the end of the data range considered (50-60 days) the estimates exhibited wide confidence intervals reflecting the stochasticity for those regions presenting small number of cases. In Figure 2(c) we presented the distribution of the first day (from the onset February 24, 2020) when the time evolution of *R_t_* converges to <1 in each region. In Table 2 the same results are listed with the corresponding calendar date together with the median *R_t_* values determined in the last 7-day time window (April, 18t^h^-24^th^).

**Table 2.**
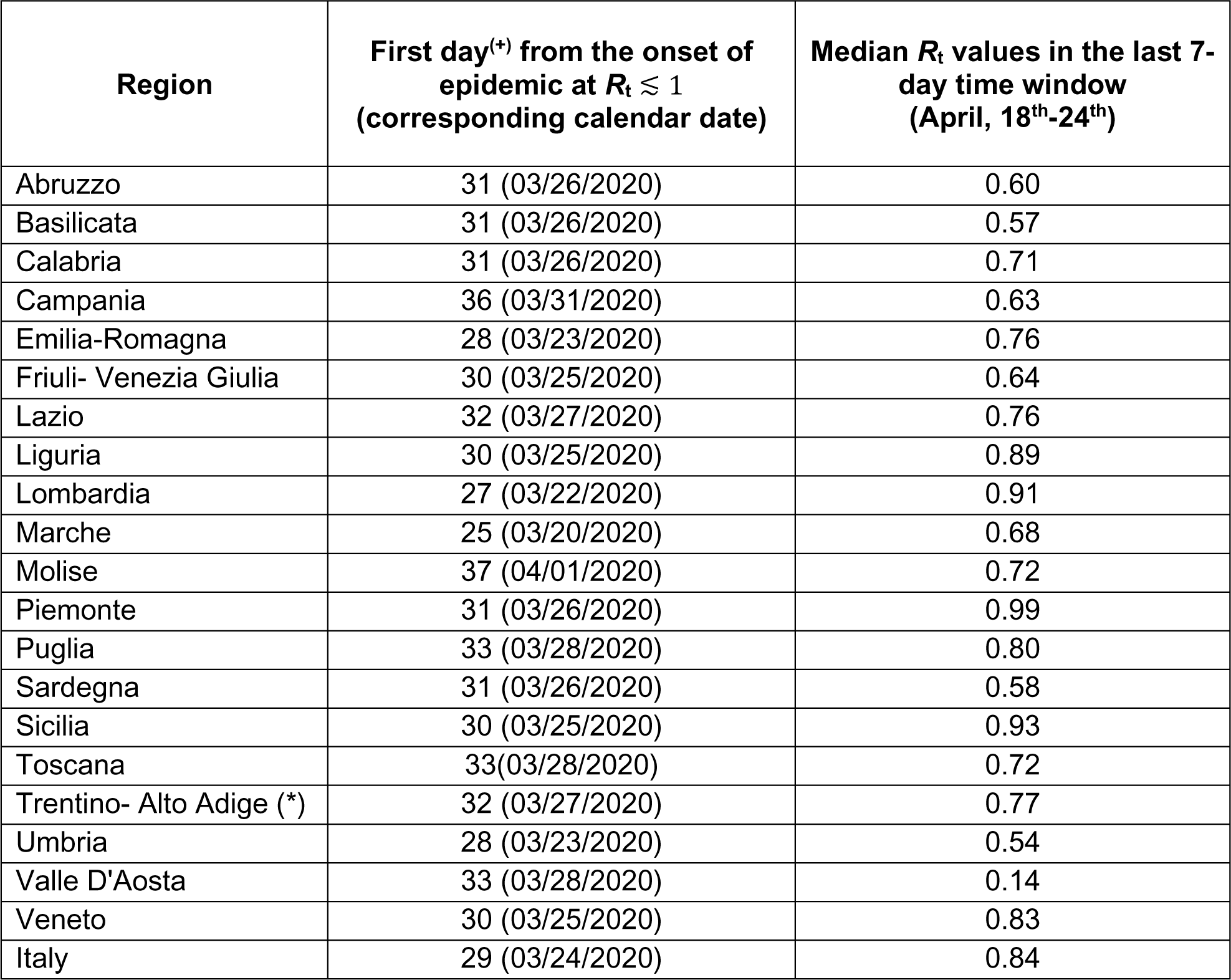
Day from the epidemic onset which *R_t_* reaches the *R* =1 condition and the median reproduction number in the last 7-day time window. (+) Date of epidemic onset February 24^th^; (*) the original incidence data related to Trento and Bolzano were merged into a single region called Trentino-Alto Adige resulting in a geographical disaggregation of Italy into 20 regions.

| Region | First day<sup>(+)</sup> from the onset of epidemic at $R_t \leq 1$<br>(corresponding calendar date) | Median $R_t$ values in the last 7-day time window<br>(April, 18<sup>th</sup>-24<sup>th</sup>) |
| --- | --- | --- |
| Abruzzo | 31 (03/26/2020) | 0.60 |
| Basilicata | 31 (03/26/2020) | 0.57 |
| Calabria | 31 (03/26/2020) | 0.71 |
| Campania | 36 (03/31/2020) | 0.63 |
| Emilia-Romagna | 28 (03/23/2020) | 0.76 |
| Friuli- Venezia Giulia | 30 (03/25/2020) | 0.64 |
| Lazio | 32 (03/27/2020) | 0.76 |
| Liguria | 30 (03/25/2020) | 0.89 |
| Lombardia | 27 (03/22/2020) | 0.91 |
| Marche | 25 (03/20/2020) | 0.68 |
| Molise | 37 (04/01/2020) | 0.72 |
| Piemonte | 31 (03/26/2020) | 0.99 |
| Puglia | 33 (03/28/2020) | 0.80 |
| Sardegna | 31 (03/26/2020) | 0.58 |
| Sicilia | 30 (03/25/2020) | 0.93 |
| Toscana | 33(03/28/2020) | 0.72 |
| Trentino- Alto Adige (*) | 32 (03/27/2020) | 0.77 |
| Umbria | 28 (03/23/2020) | 0.54 |
| Valle D'Aosta | 33 (03/28/2020) | 0.14 |
| Veneto | 30 (03/25/2020) | 0.83 |
| Italy | 29 (03/24/2020) | 0.84 |

Finally, in Figure 3, median *R_t_* values in the last 7-day time window (Table 2) are plotted in a three-dimensional array as a function of DA (Total number of active cases / km2) as recorded on April 24^th^ and population density, for all the Italian region. The array shows that similar *R_t_* are not related to similar demographic (i.e. population density) or epidemic (i.e. DA) risk factors.

**Figure 3.**
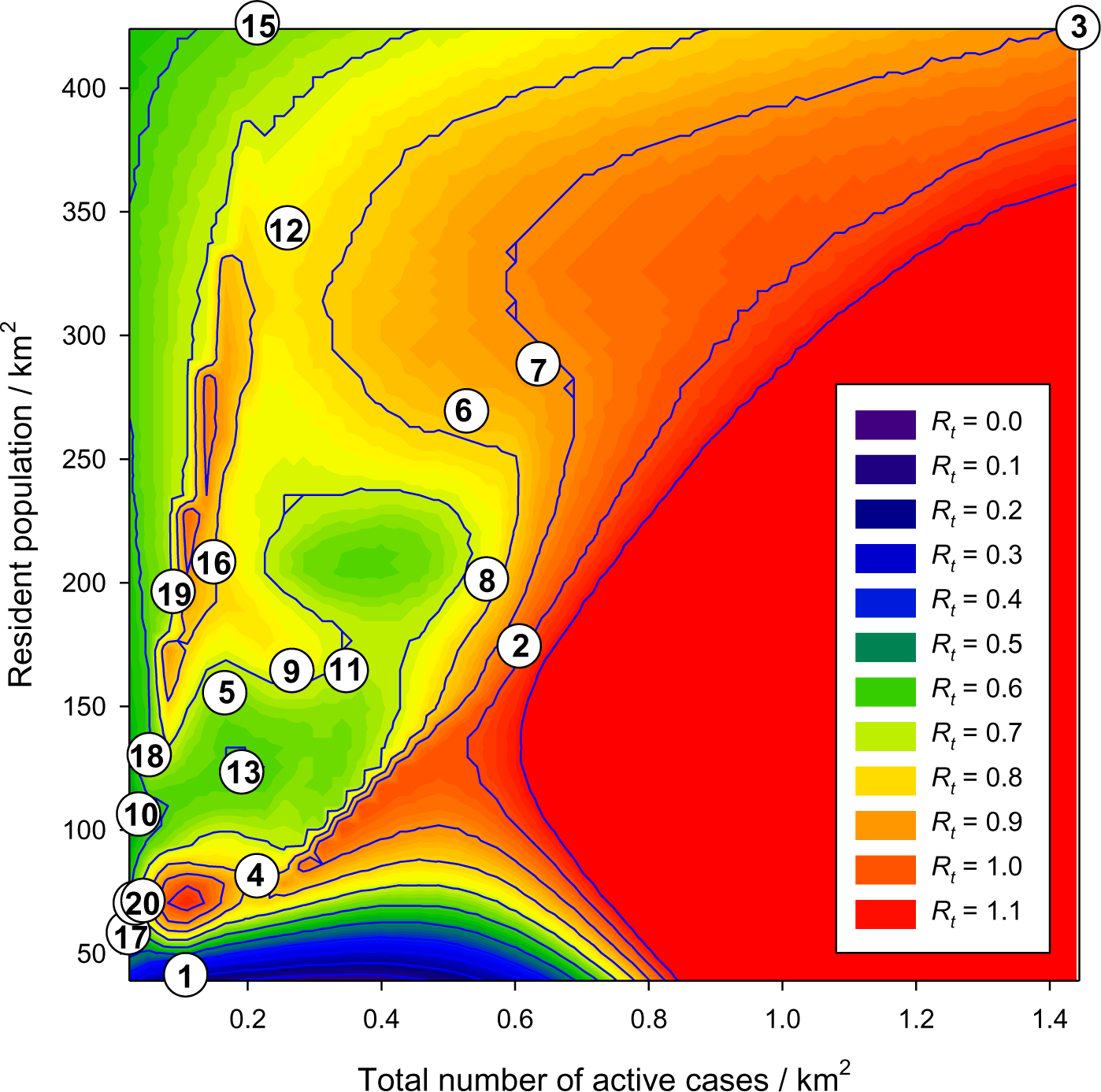
The correspondence of R, DA and population density. The false color scale represents the median R_t_ values in the period April, 18^th^ - 24^th^ as a function of DA (Total number of active cases / km^2^) as recorded on April 24^th^, and population density (Resident population / km^2^). Dots indicate data from different regions numbered as follows: 1 – Valle D'Aosta; 2 – Piemonte; 3 – Lombardia; 4 – Trentino-Alto Adige; 5 – Friuli-Venezia Giulia; 6 – Veneto; 7 – Liguria; 8 – Emilia-Romagna; 9 – Toscana; 10 – Umbria; 11 – Marche; 12 – Lazio; 13 – Abruzzo; 14 – Molise; 15 – Campania; 16 – Puglia; 17 – Basilicata; 18 – Calabria; 19 – Sicilia; 20 – Sardegna.

## 4. Discussion and conclusions

In this work, we analyzed the time evolution of incidence of the SARS-CoV-2 epidemic for two months from onset, February 24^th^ to April 24^th^, in all the Italian regions. We estimated the basic reproduction number *(R*_0_), by using the ML method in the early stage of the epidemic. In addition, we determined time evolution of this parameter across the two months of the observational period. Finally, we linked *R_t_*, with two indices, the population density and DA, the latter representing the density of infected people in a region as recorded on April 24^th^.

Firstly, we point out that these data can be considered only an approximation of the actual epidemic dynamics. Indeed, the reported number of cases strictly depends on the number of swabs that are used for Covid-19 testing and can be biased by several factors like underreporting, delays in recording as well as errors in classification of cases. [13] Therefore, large data noise is general observed, especially at the regional level, which requires a careful inspection of the epidemic curve as well as data smoothing in order to avoid unrealistic reproduction number estimation. As described in the results, for the evaluation of *R*_0_ in the initial outbreak stage, we considered data from February 24^th^, up to March 18^th^. Data in a wider range can be affected by the national lockdown on March 11^th^. This period agrees well with previous investigation where the same time window has been assumed as the infection period to determine *R*_0_ for the whole Italy.[14]

Taking these preliminary considerations into account, our result of *R*_0_ *=* 3.22 for Italy is highly consistent with values obtained by fitting the exponential growth rate of the infection across a 1-month period. [12] Similar conclusion has been drawn for Northern regions transmission dynamics and the same results were found for the Southern regions.[12] In another work, Riccardo F. et al. [2] reported *R*_0_ ranging from 2.50 to 3.00 for six selected Italian regions (Lombardia, Veneto, Emilia-Romagna, Toscana, Lazio, Puglia). Despite these values are lower than *R*_0_ obtained here, a variability of ~ 0.5 for most of the regions is thus confirmed independently of geographical location. Again, Gatto et al. [15], while including additional parameters like mobility and the spatial distribution of communities, determined a comparable initial generalized reproduction number *R*_0_ *=* 3.60. Overall, these data support the idea that epidemiological figures of the SARS-CoV-2 epidemic in Italy are slightly higher than those observed at the early stage of outbreak in Wuhan (China). [16]

The initial large values observed resulted from a sudden increase of independent first reported infections which in many cases can be related to the so called “super-spreading” events. Indeed, as observed for SARS outbreak, [9] in the early stage of the epidemic the time dependence of *R_t_* shows a fluctuating pattern characterized by wide confidence interval raised by the initial low number of cases used in the calculations. In this context, the super-spreading events cannot be necessarily triggered by a single infector, but it can be related to few people which are perpetuating an epidemic in the susceptible population.[13]

Here we observed that most of the regions have faced “super-spread events” in the early stage of epidemic. and significant is the observation of such event in southern regions Basilicata, Calabria, Campania, Molise, Puglia, Sicilia. Indeed, in these regions the overshoot of the *R_t_* observed in the first 10 days, can be likely correlated with the uncontrolled movement of people leaving the most affected northern regions to south of Italy at the beginning of March. Furthermore, it should be noted that in some regions like Valle D’Aosta and Veneto, *R_t_* sudden increases around the second week of March. Such modulations can be associated to the changes in the testing practices which promptly affected the ratio between the number of new confirmed symptomatic cases and the number of swabs owing to the step like increase of daily incidence.

After the early stages, the *R_t_* showed a decreasing trend which is likely to be affected by the temporal depletion of susceptible individuals (intrinsic factors) and by the implementation of control measures (extrinsic factors).[17] Both these factors slow down the growth rate of incidence and deeply affect the shape and time scaling of the epidemic peak driving R_t_ to fall below 1.[17]

We found that the mean value of time to reach the control regime is about 31 days from the February 24^th^ and about 14 days from the first day of nationwide lockdown (March 12^th^). This mean value is in fair agreement with the value of 29 days detected for the whole Italy proving the self-consistency of our analysis. More precisely, we noted that Marche is the first region reaching *R_t_* < 1 on March 20^th^ followed by Lombardia on March 22^nd^, Emilia-Romagna and Umbria on March 23^th^, Veneto, Friuli-Venezia Giulia, Liguria and Silicia on March 25^th^, Piemonte on March 26^th^. The last Italian region reached the control regime is the southern region Molise on 1^st^ April due to delay of the epidemic onset.

Based on the timing of the targeted (March 9^th^) and nationwide (March 12^nd^) lockdowns, this provides a direct evidence of the burden of social distancing measures introduced to control the epidemic. Indeed, the time gap between the introduction of the government measures and *R_t_* ≲ 1 is ranged between 13 and 15 days for the most affected regions namely Lombardia, Veneto, Emilia-Romagna, Piemonte. Overall, after 20 days from the national lockdown all the regions displayed reproduction number below the unity.

In the control regime, we observed for about 30 days (up to April 24^th^) that most of the regions experience a plateau in which *R_t_* fluctuates just below the unity in agreement with the smoothly decreasing of the daily incidence. Exceptions are represented by Molise and Umbria where *R_t_* drops down to ~½ % according to small number of positive cases updated. Compared to *R*_0_, the median *R_t_* of the last 7-day time window (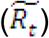) presents a quite narrow distribution with a mean value of 0.71. Considering the decrease from the mean 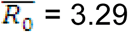 to 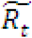 mean, we estimated that after 45 days the nationwide lockdown prevents about 78 % of potential secondary infections on average.

Although the Italian Government’s restrictive measures have proven to be of considerable utility in preventing even more devastating effects from the epidemic, the challenge in tackling “Phase 2” appears even more demanding. In this line, obtaining simple and effective indices to evaluate the state of activity of the epidemic seems mandatory: if the *R_t_* index remains essential for understanding the trend in a given area, however it is not the only parameter to account for. Briefly, if we consider two areas with the same *R_t_*, that of the two that has a population density and a higher percentage of infected people must be considered more at risk, monitored more carefully and potentially the target of more timely restrictive measures.

The population density of a given area is clearly an *R_t_*-independent risk factor for the development of an epidemic that spreads through human infection, although the population density of the different Italian regions may not be truly representative of the distribution of the population. Urban areas and in particular metropolitan areas (Rome, Milan, Naples) have a population density higher than the regional one. Furthermore, due to the peculiar Italian orography, some regions (for example Liguria, Valle D’Aosta, Trentino-Alto Adige) concentrate the population in a “habitable” area much less large than the total surface.

While admitting its arbitrariness, DA (ie the number of infected people per Km^2^) is in some way a representative parameter of how much the epidemic was active in the previous period and, above all, what is the generic risk of “meeting” a subject affection in a given area. Therefore, we suggest to associate a combined use of *R_t_* with DA and population density to evaluate the epidemic risk of a specific area, in our case of the Italian regions.

## Data Availability

The data that support the findings were derived from the following resources available in the public domain:
https://github.com/pcm-dpc/COVID-19
http://dati.istat.it/Index.aspx?QueryId=18460

https://github.com/pcm-dpc/COVID-19

http://dati.istat.it/Index.aspx?QueryId=18460

## SUPPORTING INFORMATION

**Table S1.** Demographic data, updated on January 1^st^, 2019. official data from ISTAT available at http://dati.istat.it/Index.aspx?QueryId=18460; (*): The original incidence data related to Trento and Bolzano were merged into a single region called Trentino-Alto Adige resulting in a geographical disaggregation of Italy into 20 regions.

| Region | Resident population | Surface (km<sup>2</sup>) | Population density (pers/km<sup>2</sup>) |
| --- | --- | --- | --- |
| Abruzzo | 1311580 | 10831.84 | 121 |
| Basilicata | 562869 | 10073,32 | 56 |
| Calabria | 1947131 | 15221.90 | 128 |
| Campania | 5801692 | 13670.95 | 424 |
| Emilia-Romagna | 4459477 | 22452.78 | 199 |
| Friuli- Venezia Giulia | 1215220 | 7924.36 | 153 |
| Lazio | 5879082 | 17232.29 | 341 |
| Liguria | 1550640 | 5416.21 | 286 |
| Lombardia | 10060574 | 23863.65 | 422 |
| Marche | 1525271 | 9401.38 | 162 |
| Molise | 305617 | 4460.65 | 69 |
| Piemonte | 4356406 | 25387.07 | 172 |
| Puglia | 4029053 | 19540.90 | 206 |
| Sardegna | 1639591 | 24100.02 | 68 |
| Sicilia | 4999891 | 25832.9 | 194 |
| Toscana | 3729641 | 22987.04 | 162 |
| Trentino- Alto Adige (*) | 1072276 | 13605.50 | 79 |
| Umbria | 882015 | 8464.33 | 104 |
| Valle D'Aosta | 125666 | 3260.90 | 39 |
| Veneto | 4905854 | 18345.35 | 267 |
| Italy | 60359546 | 302072.84 | 200 |

**Table S2.** Epidemiological data, updated on April 24^th^, 2020. Official data from the Dipartimento della Protezione civile available at https://github.com/pcm-dpc/COVID-19; (*) the original data related to Trento and Bolzano were merged into a single region called Trentino-Alto Adige resulting in a geographical disaggregation of Italy into 20 regions; N = aggregate number of infected people, NA = number of active infected people, DA = density of active infected people.

| Region | N<br>(persons) | NA<br>(persons) | DA<br>(persons/km<sup>2</sup>) |
| --- | --- | --- | --- |
| Abruzzo | 2803 | 2079 | 0.19 |
| Basilicata | 360 | 229 | 0.02 |
| Calabria | 1079 | 821 | 0.05 |
| Campania | 4282 | 2943 | 0.22 |
| Emilia-Romagna | 23970 | 12509 | 0.56 |
| Friuli- Venezia Giulia | 2882 | 1320 | 0.17 |
| Lazio | 6132 | 4492 | 0.26 |
| Liguria | 7173 | 3437 | 0.63 |
| Lombardia | 71256 | 34368 | 1.44 |
| Marche | 6028 | 3273 | 0.35 |
| Molise | 287 | 200 | 0.04 |
| Piemonte | 23822 | 15391 | 0.61 |
| Puglia | 3881 | 2933 | 0.15 |
| Sardegna | 1257 | 804 | 0.03 |
| Sicilia | 2981 | 2320 | 0.09 |
| Toscana | 8877 | 6133 | 0.27 |
| Trentino- Alto Adige (*) | 6232 | 2920 | 0.21 |
| Umbria | 1363 | 322 | 0.04 |
| Valle D'Aosta | 1100 | 354 | 0.11 |
| Veneto | 17229 | 9679 | 0.53 |
| Italy | 192994 | 106527 | 0.35 |

**Figure S1.**
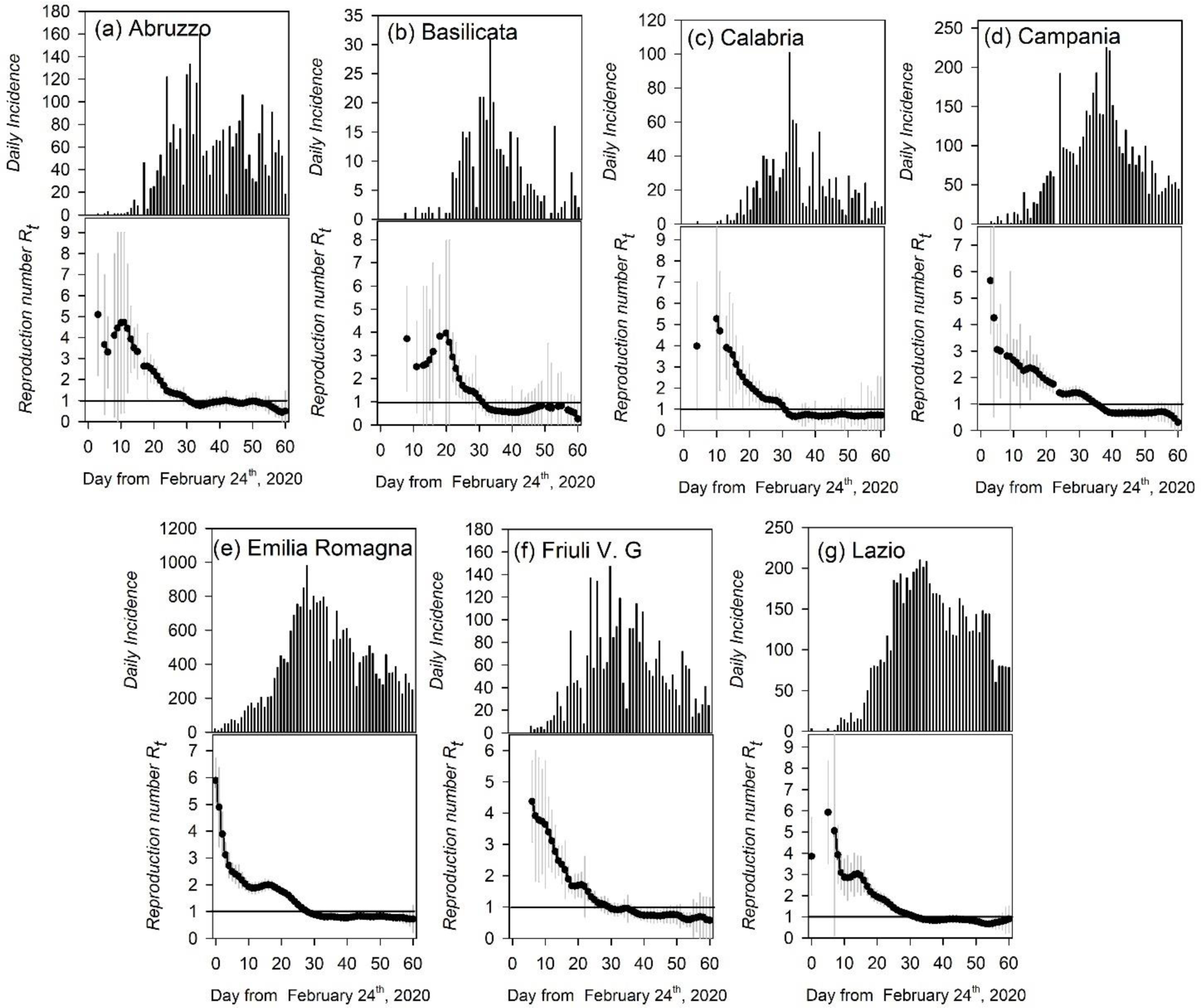
Daily incidence as numbers of new cases from February 24 to April 24 2020 for COVID-19 outbreaks (upper panel) and the corresponding time dependent reproduction number (*R_t_*) (lower panel) for the following Italian regions: (a) Abruzzo, (b) Basilicata, (c) Calabria, (d) Campania, (e) Emilia-Romagna, (f) Friuli V. G., (g) Lazio. In upper panels vertical bars are the incidence data whereas in lower panels black dots are the *R_t_* mean values accompanying by grey vertical lines standing for 95% confidence intervals. In the same panel the horizontal solid line indicates the threshold value *R* = 1, above which an epidemic will spread and below which the epidemic is controlled. Days are listed from the onset February 24^th^, 2020.

**Figure S2.**
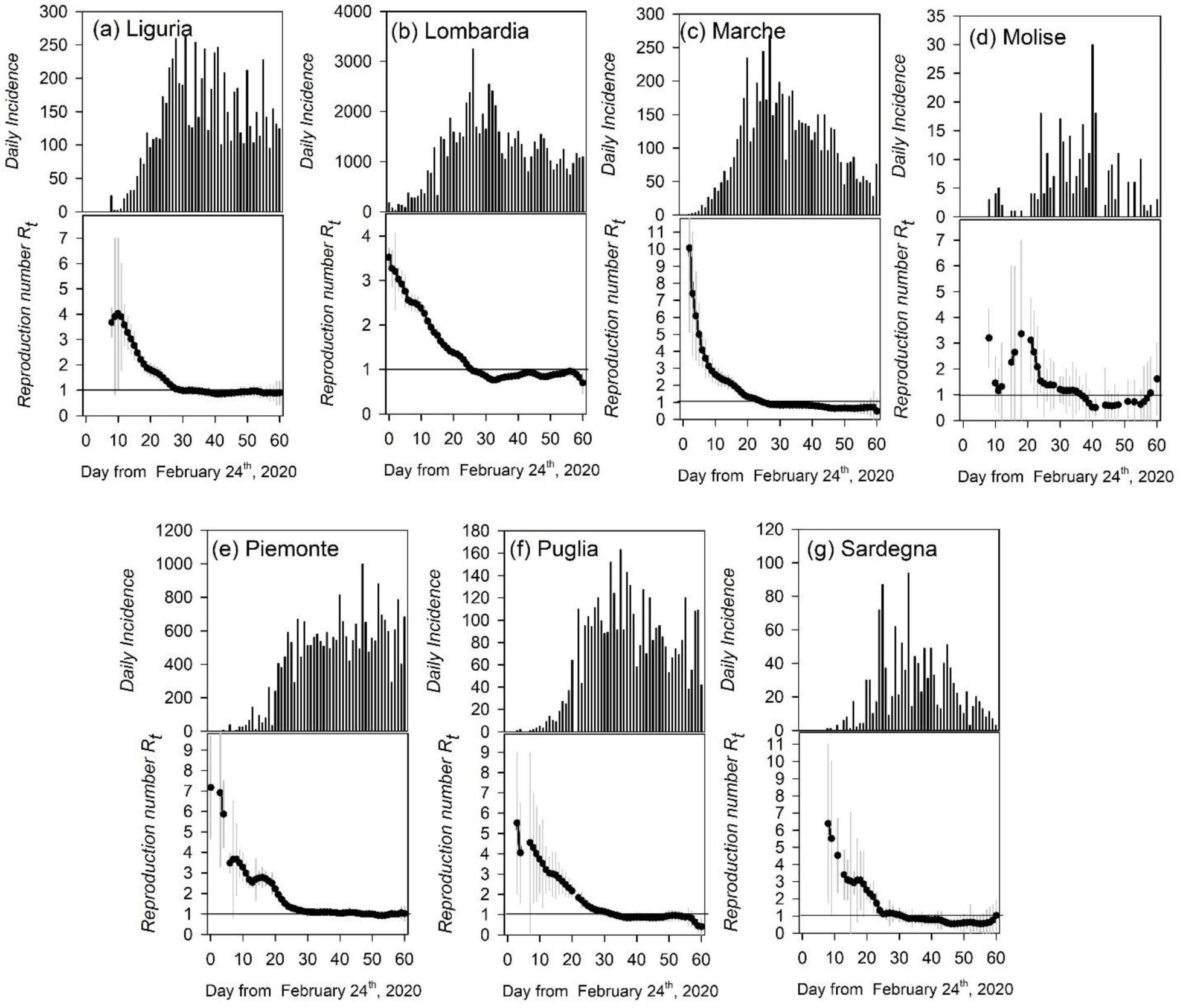
Daily incidence as numbers of new cases from February 24 to April 24 2020 for COVID-19 outbreaks (upper panel) and the corresponding time dependent reproduction number (R_t_) (lower panel) for the following Italian regions: (a) Liguria, (b) Lombardia, (c) Marche, (d) Molise, (e) Piemonte, (f) Puglia, (g) Sardegna, In upper panels vertical bars are the incidence data whereas in lower panels black dots are the *R_t_* mean values accompanying by grey vertical lines standing for 95% confidence intervals. In the same panel the horizontal solid line indicates the threshold value R = 1, above which an epidemic will spread and below which the epidemic is controlled. Days are listed from the onset February 24^th^, 2020.

**Figure S2.**
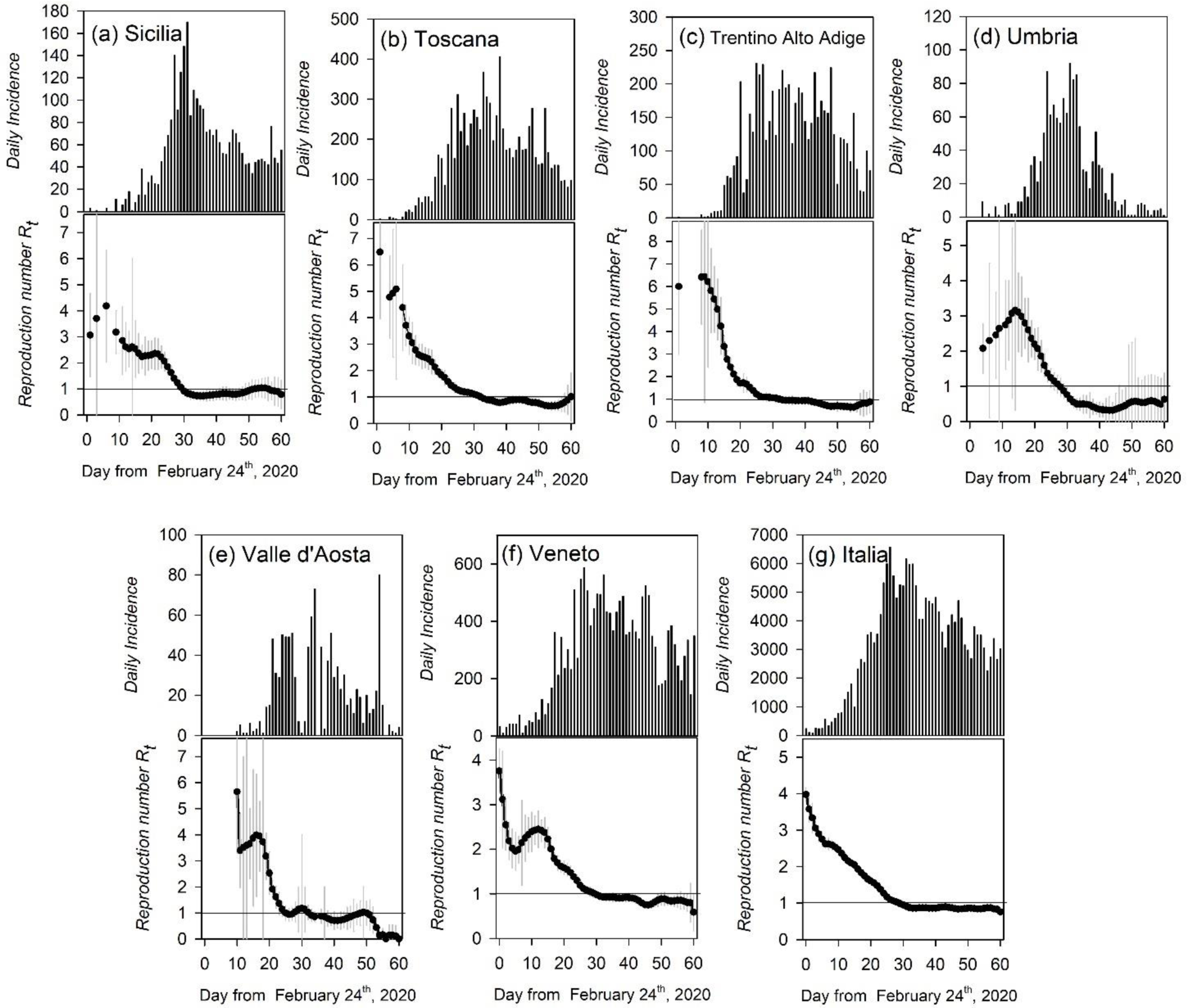
Daily incidence as numbers of new cases from February 24 to April 24 2020 for COVID-19 outbreaks (upper panel) and the corresponding time dependent reproduction number (R_t_) (lower panel) for the following Italian regions: (a) Sicilia, (b) Toscana, (c) Trentino Alto Adige, (d) Umbria, (e)Valle D’Aosta, (f) Veneto and (g) Italia. In upper panels vertical bars are the incidence data whereas in lower panels black dots are the R_t_ mean values accompanying by grey vertical lines standing for 95%% confidence intervals. In the same panel the horizontal solid line indicates the threshold value R = 1, above which an epidemic will spread and below which the epidemic is controlled. Days are listed from the onset February 24^th^, 2020.

## Notes

**Funding:** This research received no external funding

**Conflicts of Interest:** Authors declare no conflicts of interest

### Competing Interest Statement

The authors have declared no competing interest.

### Funding Statement

No external funding was received

